# Isolation of SARS-CoV-2 from the air in a car driven by a COVID patient with mild illness

**DOI:** 10.1101/2021.01.12.21249603

**Authors:** John A. Lednicky, Michael Lauzardo, Md. M. Alam, Maha A. Elbadry, Caroline J. Stephenson, Julia C.Gibson, J. Glenn Morris

## Abstract

We used a Sioutas personal cascade impactor sampler (PCIS) to screen for SARS-CoV-2 in a car driven by a COVID-19 patient. SARS-CoV-2 was detectable at all PCIS stages by PCR and was cultured from the section of the sampler collecting particles in the 0.25 to 0.50 □μm size range.

---

While we have learned a great deal about transmission of SARS-CoV-2 since the beginning of the current pandemic, questions remain about the exposure risk in different settings, and the contribution of various modes of transmission to virus spread.^1^ In particular, there is continuing uncertainty about the relative contribution of larger virus-laden respiratory particles at close distances, compared with virions in aerosols at close or longer ranges, to spread of the virus. There are now multiple epidemiology studies consistent with aerosol spread of SARS-CoV-2 within closed spaces,^2,3^ and the virus has been shown to remain infective in laboratory-generated aerosols for at least 16 hours.^4^ Data from our group and others have documented the presence of the virus in aerosols in patient settings by RT-PCR.^5-7^ However, molecular detection of SARS-CoV-2 RNA does not necessarily correlate with risk of developing COVID-19, since only viable virions can cause disease. Subsequently, we have reported isolation of viable SARS-CoV-2 from the air within the room of hospitalized COVID-19 patients.^8^

The current study was undertaken to assess SARS-CoV-2 transmission in a “real life” setting, outside of a medical facility. In epidemiologic studies, public transportation vehicles have been identified as a risk factor for transmission by CDC, and studies from Singapore have reported an odds ratio for infection of 3.07 [95% CI 1.55-6.08] for persons sharing a vehicle with an infected person.^9^ In this setting, we were interested in documenting that viable virus could be isolated from the air of a car being driven by a person infected with SARS-CoV-2. We were also interested in determining the size distribution of airborne respiratory secretions that contain virions to provide some guide to the contribution of droplets and aerosols to transmission risk.

To do this, we asked a COVID patient with minimal symptoms to drive her car for a short period of time with a Sioutas personal cascade impactor sampler (PCIS)^10^ clipped onto the sun-visor above the passenger seat next to her. The Sioutas PCIS separates airborne particles in a cascading fashion and simultaneously collects the size-fractionated particles by impaction on polytetrafluoroethylene (PTFE) filters. It has collection filters on four impaction stages (A–D), and an optional after-filter can be added onto a 5th stage (E). The PCIS separates and collects airborne particulate matter above the cut-point of five size ranges: >2.5□μm (Stage A), 1.0 to 2.5□μm (Stage B), 0.50 to 1.0□μm (Stage C), 0.25 to 0.50□μm (Stage D), and <0.25□μm (collected on an after-filter) (Figure 1); details of the instrumentation and laboratory methods are included in supplementary material. For the current study, a PCIS (SKC, Inc., catalog number 225-370) unit was used with a Leland Legacy pump (SKC, Inc., cat number 100-3002) and operated at a flow rate of 9□L/min.

**Figure 1.**
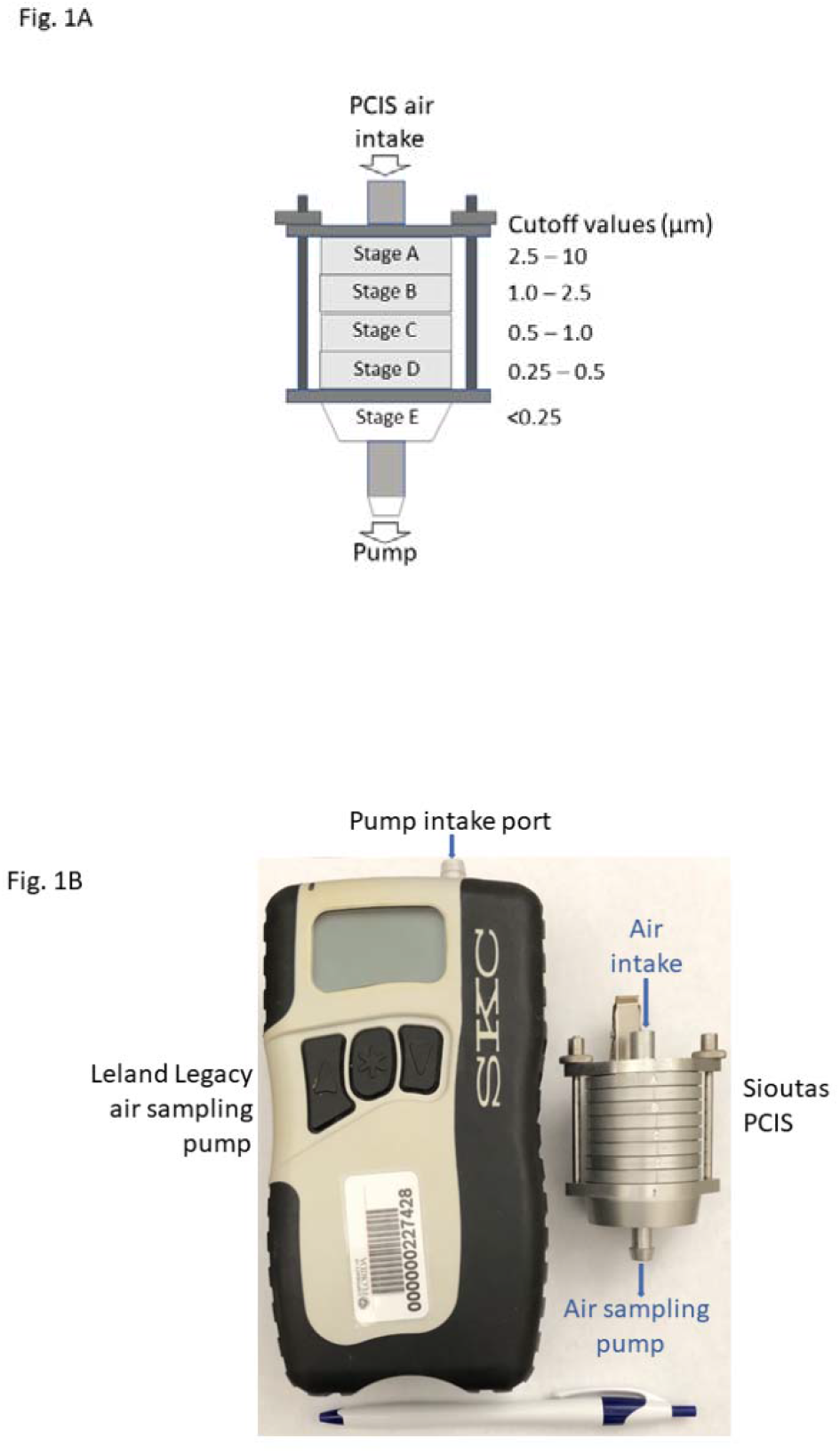
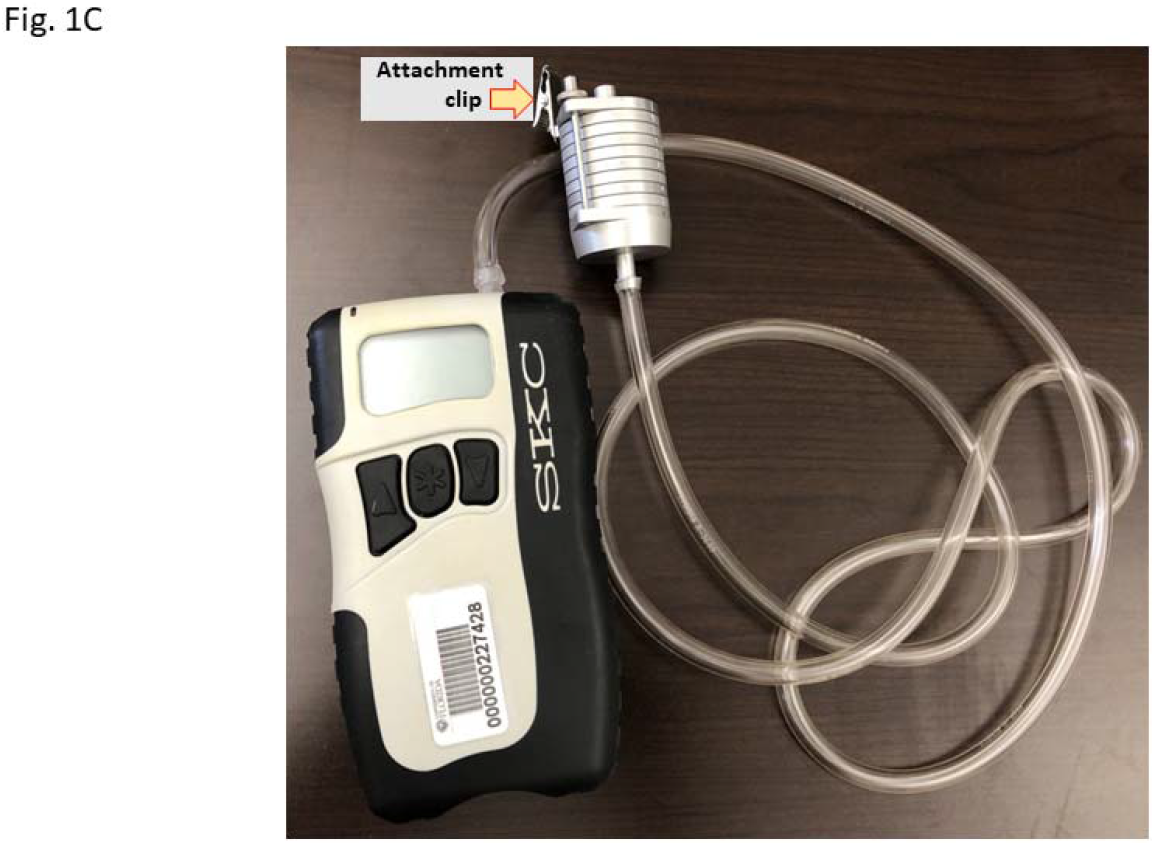
Sioutas Personal Cascade Impactor Sampler (PCIS). (A) Schematic representation of the five stages of a PCIS. (b) Photograph of an SKC Leland Legacy air sampler pump and a PCIS unit. A pen is shown below the pump and PCIS unit to provide size perspective. (c) Attachment clip in PCIS and SKC Leland Legacy air sampler pump assembly.

The patient, who was in her twenties, had initially presented to clinic with a one-day history of mild fatigue, nasal congestion, and sore throat, following exposure to a roommate who had a laboratory-confirmed diagnosis of COVID-19. The patient denied fever, cough, shortness of breath, or other symptoms, and had a normal physical exam. In testing performed by the UFHealth clinical laboratory, a nasopharyngeal swab collected at the time of presentation was positive by RT-PCR. The patient was advised to isolate at home for a 10-day period of time, and appropriate contact tracing was initiated.

Two days after the diagnostic sample was obtained, the patient agreed to have the PCIS placed on the passenger seat of her car (an older model Honda Accord) for the drive from the clinic to her home. Her symptoms at that time were minimal, with no cough. The PCIS was attached to the sun-visor on the passenger side of the car, approximately 3 feet from the patient’s face and with the intake port pointing toward the roof of the car. During the 15-minute drive the patient was not wearing a mask. The air conditioner in the car was on: during the drive the temperature within the car’s cabin ranged from 24.2 to 22.8 °C and relative humidity fluctuated from 55.2 to 42.5%; outside temperature was 32 °C and relative humidity 99%.

Two hours after the patient’s arrival home, an investigator wearing personal protective equipment (N95 mask, gloves, and a Tyvek laboratory coat) opened the car and turned off the pump of the air sampler, and transferred the PCIS-pump assembly into a sealed container and decontaminated the outer surface of the container; the calculated amount of air filtered from inside the vehicle was approximately 1.2□m^3^. After removal from the car, the air sampling assembly was transported with appropriate biosafety precautions to a BSL2-enhanced laboratory at the University of Florida. Within 30 minutes of the termination of air-sampling, the PCIS filters were individually immersed in 1 ml of recovery solution (PBS with 0.5%□w/v BSA fraction V and 0.2 M surcose^11^) for 30 minutes at room temperature, and then scraped with flocked swabs pre-wetted with recovery solution. Samples were concentrated by centrifugation and stored at -80°C.

After one thaw on ice, RNA was extracted from 140 µl of material extruded off the PCIS filters using a QIAamp Viral RNA Mini Kit. rRT-PCR to detect SARS-CoV-2 RNA was performed using primer system Led-N-F, Led-N-R, and Led-N-probe.^11^ The number of viral genome equivalents present in each sample was estimated from the measured quantification cycle (Cq) values, and was attained using 10-fold dilutions of calibrated plasmids containing an insert of the SARS-CoV-2 N-gene as previously described.^8^ SARS-CoV-2 RNA was detected on filters A to D, suggesting that the PCIS had collected a range of particle sizes containing SARS-CoV-2 virions or other material (possibly cell debris) containing SARS-CoV-2 RNA (Table 1). More of the SARS-CoV-2 RNA material was collected on filter D than that on filters A–C and E combined.

**Table 1.** rRT-PCR Detection of SARS-CoV-2 RNA on filters.

| **Sample** | **PCIS size cut-off value (µm)** | **Cq** | **Genome eq. 25 µl rRT-PCR reaction<sup>a</sup>** | **Genome eq. per 1.0 m<sup>3</sup> of air** |
| --- | --- | --- | --- | --- |
| PCIS filter A | 2.5 – 10.0 | 36.66 | 3.77E+01 | 1.24E+03 |
| PCIS filter B | 1.0 – 2.5 | 35.23 | 1.19E+02 | 3.90E+03 |
| PCIS filter C | 0.5 – 1.0 | 34.37 | 2.37E+02 | 7.77E+03 |
| PCIS filter D | 0.25 – 0.5 | 33.50 | 4.79E+02 | 3.14E+04 |
| PCIS filter E | < 0.25 | 40.1 | (equivocal) | 0 |
| + control | Not applicable | 26.85 | 1.00E+05 | Not applicable |
| - control | Not Applicable | 0 | 0 | Not Applicable |

Standard curve: SYBR, E=123.4%, R<sup>2</sup>=1, slope = -2.865, Y int = 41.173
<sup>a</sup>Equivalent to RNA in 10 µl out of a total resuspension volume of 400 µl/filter.

Filter material was subsequently inoculated onto Vero E6 cells as previously described.^8^ Early cytopathic effects consistent with those caused by SARS-CoV-2 were observable by 3 days in cells inoculated with material collected onto PCIS filter D; by day 5, foci of infection were apparent for cells inoculated with material from filter D (Supplemental Figure 1C and D), with no signs of virus infection in cells inoculated with material collected by PCIS filters B, C, and E. The mock-infected cell monolayer remained intact (Supplemental Figure 1A), fungus overgrowth was evident in cells inoculated with material from PCIS filter A (Supplemental Figure 1B), and rRT-PCR tests indicated that SARS-CoV-2 had indeed been isolated. For further confirmation, an aliquot (20 µl) of the virus collected 5 days post inoculation of material from filter D was passaged in Vero E6 cells, wherein an rRT-PCR Cq value of 12.46 was attained 3 days post-inoculation of the cells.

Sanger sequencing was performed on RNA extracted from the cell growth medium corresponding to PCID filter D; as a secondary check, NGS was also performed using an Illumina MiSeq platform.^8^ Sequencing reactions were performed at different locations by different personnel. The virus isolated was designated as SARS-CoV-2/human/USA/UF-29/2020 (GenBank no. MW229264.1). SARS-CoV-2 UF-29 has the following marker variants relative to reference strain Wuhan-Hu-1 (GenBank no. NC_045512.2): C241T, C3037T, A23403G, G25563T, Spike protein D614G, and ORF3a protein Q57H. These markers identify it as a member of clade GH following Global Initiative on Sharing All Influenza Data (GISAID) clade nomenclature.

The air we breathe typically contains airborne particles of biological origin, including bacteria (cells and spores), fungi (mycelia and spores), and virions. Respiratory pathogens that are able to remain viable after aerosolization and air transport are potential causes of respiratory diseases, and they are often associated with other substances to form ‘complex particles.’^11-12^ It should be noted that scanning electron microscopy of human specimens that contain SARS-CoV-2 often depict clumps of virions and virions attached in a beads on a string manner (example: see https://www.flickr.com/photos/54591706@N02/49557785797). Therefore, it is plausible that humans release different forms of the virus (clumped, single particle, etc.), which affects how the virions are partitioned in different-sized airborne respiratory secretions. Our detection of vRNA in stages A to D of the impactor is consistent with the hypothesis that virions are present in different sized respiratory secretions. Remarkably, viable virus was only detected in stage D, corresponding to collection of airborne particles of the size range of 0.25 to 0.5 µm.

The PCIS is a cascade impactor, and the manner that virus particles are collected (impaction onto filters) and the presence of a constant air-flow stream (drying out the virus) should, if anything, have reduced virus viability. The air flow in the impactor increases in velocity as it passes through stages A to E, imperiling virions collected at stage D to a greater extent than virions collected at earlier stages. Yet despite these potential adverse conditions, we isolated viable virus from stage D of the impactor. Taken together, our data highlight the potential risk of SARS-CoV-2 transmission by minimally symptomatic persons in the closed space inside of a car (with closed windows and air conditioning running), and suggest that a substantial component of that risk is via aerosolized virus.

## Data Availability

Sequence data have been deposited in GenBank. All other data are presented in the manuscript

## Lednicky *et al*: Supplemental Material

### Details regarding instrumentation and laboratory methods

#### Instrumentation

The Sioutas Personal Cascade impactor sampler (PCIS) separates airborne particles in a cascading fashion and simultaneously collects the size-fractionated particles by impaction on polytetrafluoroethylene (PTFE) filters.^1,2^ It has collection filters on four impaction stages (A–D), and an optional after-filter can be added onto a 5th stage (E). The PCIS separates and collects airborne particulate matter above the cut-point of five size ranges: >2.5□μm (Stage A), 1.0 to 2.5□μm (Stage B), 0.50 to 1.0□μm (Stage C), 0.25 to 0.50□μm (Stage D), and <0.25□μm (collected on an after-filter) (Figure 1). PTFE filters (Teflon filters) can collect particles at high efficiency above the cut-points without the need for coatings,^1^ and that is advantageous, as various coatings reduce the recovery efficiency of viable virus. For the collection of airborne viruses, the filters are not prewetted with methanol prior to use, which helps preserve virus viability.^3^ For the current study, a PCIS (SKC, Inc., catalog number 225-370) unit was used with a Leland Legacy pump (SKC, Inc., cat number 100-3002) and operated at a flow rate of 9□L/min. PTFE filters (25□mm, 0.5□μm pore, SKC, Inc. catalog number 225-2708) were used for the collection stages A to D, and a PTFE after-filter (37□mm, 2.0□μm pore, SKC Inc., catalog number 225-1709) for stage E. The pump’s operating flow rate (9 L/min) was calibrated by measuring volume displacement using a Defender Primary Standard Calibrator (SKC, Inc., catalog number 717-510H). The patient drove the car for 15 minutes with windows closed and the air-conditioner on, then parked it and exited, leaving the windows up and the air sampler running for an additional 120 minutes. A total collection time of 135□min was thus used to sample approximately 1.22□m^3^ of air within the vehicle.

### Laboratory Methods

Within 30 minutes of the termination of air-sampling, the PCIS filters were individually immersed in 1 ml of recovery solution (PBS with 0.5%□w/v BSA fraction V and 0.2 M surcose)^4^ for 30 minutes at room temperature to help rehydrate and dislodge virions stuck on the filter surfaces. The filters and fluid were then transferred to a sterile plastic petri dish, and the filters scraped with flocked swabs pre-wetted with recovery solution and residual fluid in each swab extruded into the liquid corresponding to each filter. The recovery solutions were concentrated by centrifugation in Amicon Ultra-15 centrifugal filter units with Ultracel-100 membranes with a molecular mass cutoff of 100 kDa (Millipore, Bedford, MA) at 4,000 × g for 12 min to a volume of approximately < 400 μL, and the concentrates adjusted to 400 µL by addition of recovery solution. They were then aseptically transferred to sterile plastic cryotubes with O-ring seals, and the tubes stored in a locked -80°C freezer for subsequent analyses.

After one thaw on ice, RNA was extracted from 140 µl of material extruded off the PCIS filters using a QIAamp Viral RNA Mini Kit and the purified bound RNA eluted from the RNA-binding silicon columns in a volume of 80 µl. rRT-PCR was subsequently used to detect SARS-CoV-2 RNA and was performed using primer system Led-N-F, Led-N-R, and Led-N-probe.^4^ Briefly, vRNA was denatured for 5□min at 67°C in the presence of SUPERase-In RNase inhibitor (Invitrogen Corp.), cooled rapidly, and 25 µl rtRT-PCR performed in a BioRad CFX96 Touch Real-Time PCR Detection System using 5 μL of purified vRNA using the following parameters: 400 nM final concentration of forward and reverse primers and 100 nM final concentration of probe using SuperScript™ III One-Step RT-PCR system with Platinum™ Taq DNA Polymerase (Thermo Fisher Scientific). Cycling conditions were 20 minute at 50 °C for reverse transcription step, followed by 2 minutes at 95 °C for Taq polymerase activation step, then 44 cycles of 15 seconds at 95 °C of denaturing, 30 seconds at 57 °C for annealing, and 20 seconds at 68 °C for extension.^5^ The number of viral genome equivalents present in each sample was estimated from the measured quantification cycle (Cq) values, and was attained using 10-fold dilutions of calibrated plasmids containing an insert of the SARS-CoV-2 N-gene as previously described.^4^

Attempts to isolate SARS-CoV-2 were performed in a BSL3 laboratory by trained analysts wearing full head-covering powered air purifying respirators and appropriate PPE. After one thaw on ice, 150□μL aliquots of PCIS fluids were inoculated onto newly confluent Vero E6 cells, which were then incubated at 37 °C in a humidified 5% CO_2_ incubator. Mock infected cells were maintained in parallel. The cells were re-fed with cell growth medium containing reduced serum (3% fetal bovine serum) every three days.^4^

Early cytopathic effects consistent with those caused by SARS-CoV-2 were observable by 3 days in cells inoculated with material collected onto PCIS filter D; by day 5, foci of infection were apparent for cells inoculated with material from filter D (Supplemental Figure 1C and D), with no signs of virus infection in cells inoculated with material collected by PCIS filters B, C, and E. The mock-infected cell monolayer remained intact (Supplemental Figure 1A), fungus overgrowth was evident in cells inoculated with material from PCIS filter A (Supplemental Figure 1B), and rRT-PCR tests indicated that SARS-CoV-2 had indeed been isolated. For further confirmation, an aliquot (20 µl) of the virus collected 5 days post inoculation of material from filter D was passaged in Vero E6 cells, wherein an rRT-PCR Cq value of 12.46 was attained 3 days post-inoculation of the cells.

Sanger sequencing was performed on RNA extracted from the cell growth medium corresponding to PCID filter D. As a secondary check, NGS was also performed using an Illumina MiSeq platform, as previously described.^4^ The sequencing reactions were performed at different locations by different personnel.

